# Geometry of navigation identifies genetic-risk and clinical Alzheimer’s disease

**DOI:** 10.1101/2023.10.01.23296035

**Authors:** Uzu Lim, Rodrigo Leal Cervantes, Gillian Coughlan, Renaud Lambiotte, Hugo J. Spiers, Michael Hornberger, Heather A. Harrington

## Abstract

Recent research evidence demonstrates that the inability to orient oneself and navigate space is an early indicator of Alzheimer’s Disease. The video game Sea Hero Quest (SHQ) was designed to assess the players’ navigation ability, and several research works analysed the SHQ data using simple metrics such as length and time of navigation paths. Expanding these analyses, we propose new performance metrics that capture the geometry of paths, and analyse datasets of more than 60,000 navigators. The metrics identify players who failed the navigation task, the dementia patients, and carriers of the at-risk allele of the Apolipoprotein-E [APOE]. Furthermore the metrics detect weak navigation ability when only a fraction of navigation paths are used, with superior performance to baseline methods. Our findings demonstrate that the proposed performance metrics pave the way to a comprehensive pre-clinical screening toolbox for Alzheimer’s Disease.

**TEASER:** We propose geometric methods to capture decline in navigation ability from dementia.

## INTRODUCTION

Much recent research evidence indicates that a decline of spatial navigation ability precedes the onset of Alzheimer’s disease (AD) (Bierbrauer et al., 2020; Coughlan et al., 2018, 2019; Kunz et al., 2015). Problems with spatial navigation could become apparent before episodic memory deficits, which are currently used as the gold standard for AD diagnosis (McKhann et al., 2011). Navigation ability, however, varies naturally from individual to individual and it is influenced by environmental factors (Coutrot et al., 2022). To establish benchmark data for spatial navigation to establish new metrics and thresholds for diagnostics and disease monitoring in AD, the video game Sea Hero Quest (SHQ) was developed. SHQ has been downloaded more than 4 million times on mobile devices, and a subset of the players were stratified using demographic information including country of residence, age, and gender (Coutrot et al., 2018; Spiers et al., 2021). The game gives various navigation tasks to its players, consisting of different numbered levels, where each level consists of a navigation task to reach physical landmarks called checkpoints (Figure 1). Data collected on SHQ provides a benchmark to determine what is the natural variability that can be expected given the demographic characteristics of an individual. Navigation ability in the virtual terrain of SHQ was shown to have a significant correlation with navigation ability in the real world (Coutrot et al., 2019).

**Figure 1.**
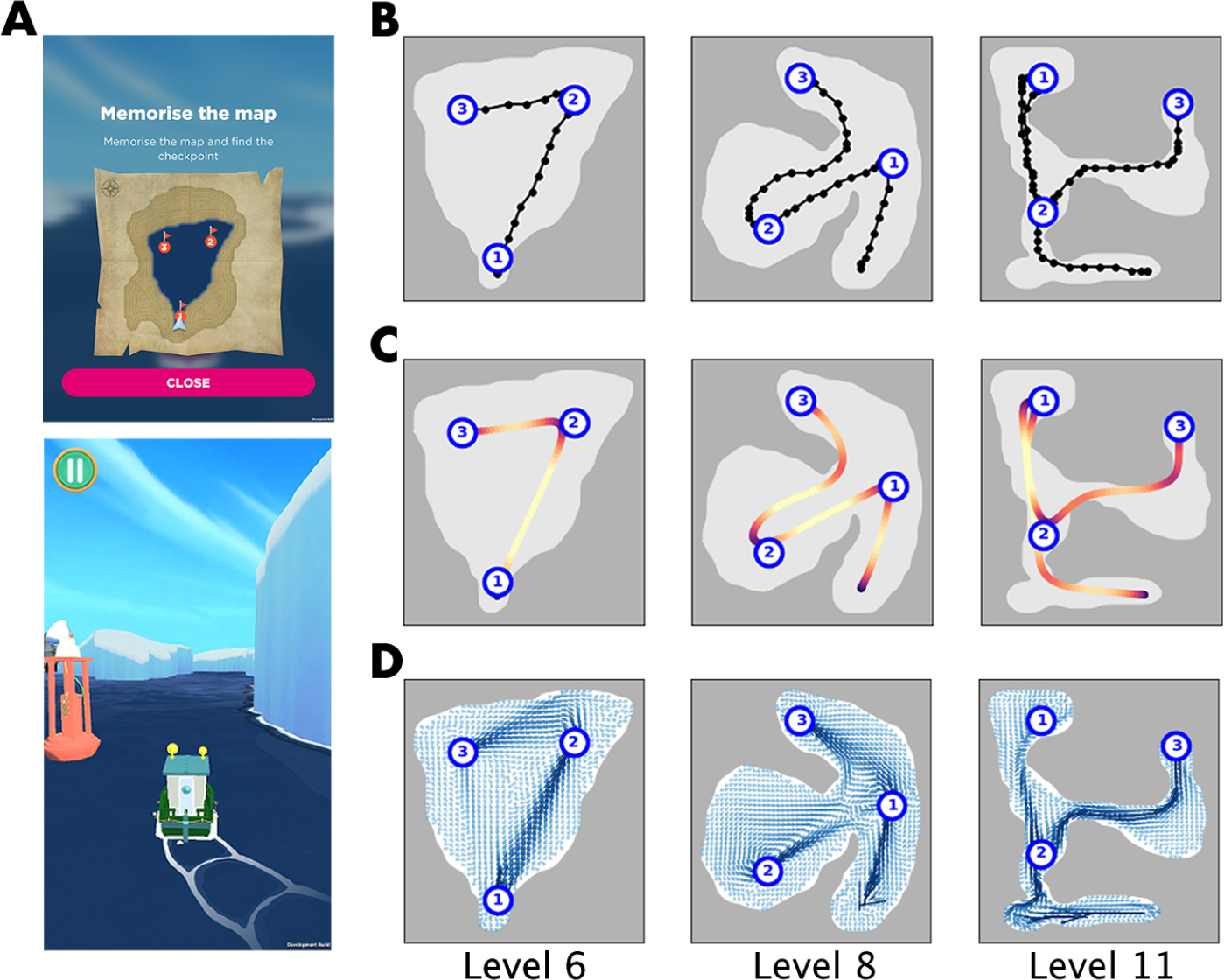
Path navigation challenges in the mobile game Sea Hero Quest (SHQ), and visualisations of gameplay data associated with three gameplay Levels: 6, 8, and 11. (A): Scenes from a SHQ gameplay, in Level 6. Top: a map with numbered checkpoints that the players must visit in the correct order. Bottom: the player then must navigate the terrains without looking at the map they were provided. (B): Efficient navigation paths through Levels 6, 8, 11 (from left to right). The dots indicate the original low-resolution navigation data, with highly discretized map coordinates (approximately 50 x 50) recorded in the frequency of 2Hz. (C): The same navigation paths in (B) with higher resolutions, reconstructed to 3 times the frequency using Gaussian kernel smoothing. The colour on the path indicates the speed of navigation, with dark purple indicating low speed and bright yellow indicating high speed. (D.) Population average behaviour of gameplay, where small arrows are drawn towards the average direction each player took at each map coordinate. The arrows are called the mobility field, and it is used to compute V-Conformity, a particularly effective metric out of the 6 proposed metrics.

Previous research demonstrated that the SHQ game provides a means of distinguishing healthy ageing adults from adults genetically at-risk of AD (defined by apolipoprotein E [*APOE*] ε4 allele carriership status) (Coughlan et al., 2019, Coughlan et al., 2020a, Coughlan et al., 2020b, Coughlan et al., 2021). The benchmark dataset allowed to determine at-genetic-risk status on an individual level, using two performance metrics: path length and duration in the SHQ game. These simple measures are clinically viable and easily implemented in diagnostic setups, but they do not capture the full richness of the navigation trajectory data.

The current study overcomes this limitation by proposing 6 performance metrics that capture geometric properties of navigation, in addition to length and duration of paths. The performance metrics were chosen on the basis of our previous findings (Coughlan et al. 2019, Kunz et al. 2015, Bierbrauer et al. 2020), specifically that suboptimal navigation in SHQ will (1) Take longer to complete, (2) Curve around unnecessarily, (3) Steer towards the boundary^1^, and (4) Deviate from the average behaviour of other players. The total of 8 performance metrics used in this study correspond to (1)-(4) as: (1) Duration, Length, (2) Curvature, (3) Boundary affinity, (4) F-Deviation, S-Deviation, M-Conformity, V-Conformity. (See Box 1 for details). These metrics together provide a more comprehensive way to understand spatio-temporal navigation paths, rather than to rely on only length (a spatial measure) and duration (a temporal measure) of navigation.

Two types of datasets were used to evaluate efficacy of the proposed metrics: (1) Benchmark dataset: a large dataset (N = 60,976) players in the United Kingdom without any clinical information (Coughlan et al., 2019), and (2) Labelled dataset: a smaller dataset (N = 77) with biological labels consisting of 3 cohorts: (2A) players with a clinical diagnosis of Alzheimer’s Disease, (2B) cognitively unimpaired APOEε3ε3 players, and (2C) cognitively unimpaired APOEε3ε4 players without AD. We test the efficacy of proposed metrics on the large Benchmark dataset by measuring each metric’s capability to distinguish two player groups: players that visit checkpoints correctly vs. incorrectly. In the smaller Labelled dataset, we measure performance of players with AD, APOEε3ε3 allele, and APOEε3ε4 allele, relative to the large Benchmark dataset, and study the distribution of percentile ranks. Finally, we assess capabilities of metrics in early prediction of navigation ability, by testing how partial segments of navigation paths can still distinguish players that visit checkpoints correctly.

### Box 1.

Performance metrics for assessing navigation

The following are the simplified descriptions of the performance metrics. The metrics 1, 2 were already studied extensively, and the metrics 3 to 8 are proposed in this study. See the Methods section for mathematically precise definitions.

1. **Length:** Length of the navigation path.
2. **Duration:** Duration taken for the navigation to complete a level.
3. **Curvature:** Average curviness of a navigation path. Defined as the sum of the rate of change of the direction of movement, averaged across navigation path length.
4. **Boundary affinity:** Average closeness to the map boundary. Defined as the sum of a sigmoid function value measuring distance from the map boundary, averaged across navigation path length.
5. **F-Deviation:** Deviation from benchmark navigation behaviour. Defined as the Frobenius matrix norm of the difference between two matrices A-B, where A describes the population behaviour and the B describes the individual behaviour to be assessed.
6. **S-Deviation:** A variant of F-deviation using the supremum matrix norm instead of the Frobenius matrix norm.
7. **M-Conformity:** Average conformity to the benchmark navigation behaviour. Defined as the weighted sum of matrix entries describing frequency of movements along map coordinates, where the weight is given by the frequency of the movement in the benchmark population. M stands for “matrix”.
8. **V-Conformity:** Average conformity to the benchmark navigation behaviour. Defined as the sum of the dot-product between the navigation path’s velocities and that of the benchmark population. V stands for “vector”.

## RESULTS

### (1) Validation: metrics capture unique aspects of navigation behaviour

Using the proposed performance metrics (Box 1), we first study the qualitative nature of the performance metrics by tabulating the best-performers and the worst-performers according to each performance metric (Figure 2). The top 3 rows and bottom 3 rows of Figure 2 respectively correspond to best and worst navigation paths, according to each of the 8 performance metrics. The strong contrast between the top and bottom rows serves as a direct validation that *each* metric is able to distinguish efficient and inefficient navigations. Furthermore, we observe a uniformity among efficient navigation and a diversity among inefficient navigation; different metrics penalise different traits of navigation. We also find that pairwise correlation coefficients of metrics are positive, with the average value of 0.605, thus showing both consistency and diversity of behaviours captured (Figure 3).

**Figure 2.**
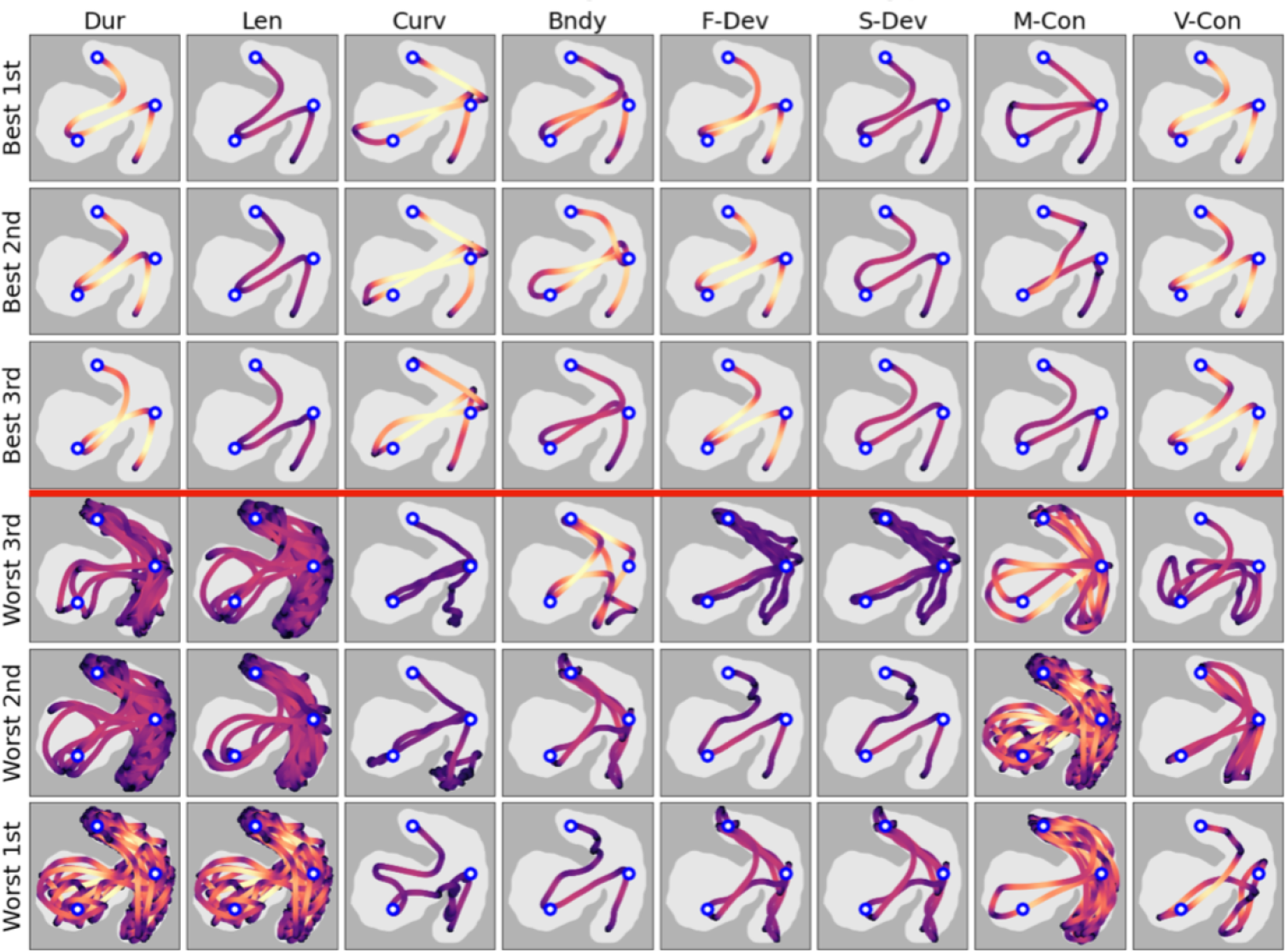
The best and worst navigation paths in Level 8, according to the 8 performance metrics. Each row corresponds to a performance metric and the columns show the top-scoring and bottom-scoring navigation paths according to each performance metric. Each navigation path has been coloured according to speed at each point, where dark purple corresponds to low speed and bright yellow corresponds to high speed.

**Figure 3.**
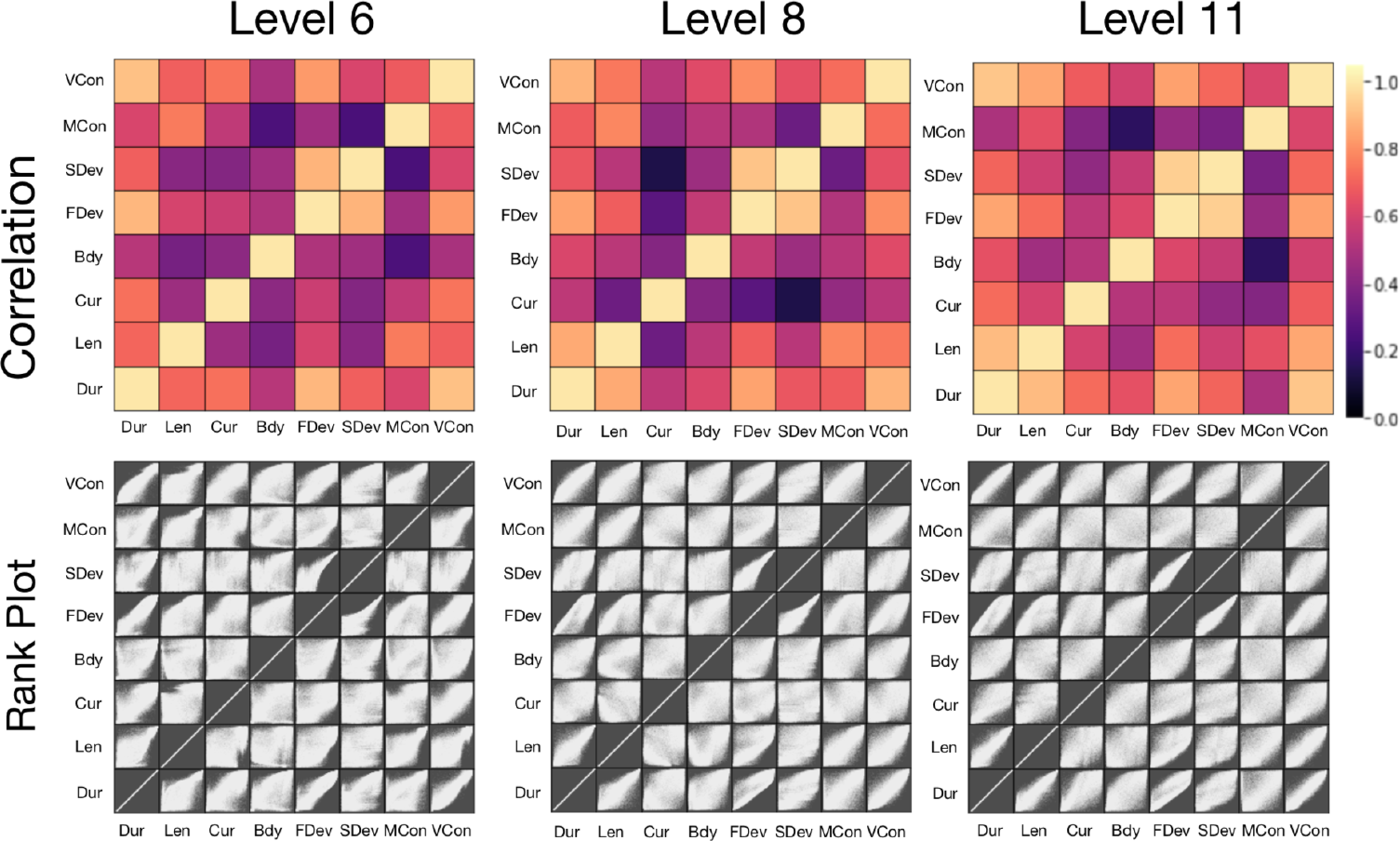
Correlation plots and rank plots comparing the 8 performance metrics, across Levels 6, 8, and 11. In each correlation plot (top row), we have a 8 x 8 tabulation of pairwise comparison between metrics. Dark purple indicates low correlation, and bright yellow indicates high correlation. In each rank plot (bottom row), we again have a 8 x 8 tabulation of pairwise comparisons. In each small square in the 8 x 8 tabulation, a scatterplot of percentile ranks is shown. Each navigation path is given 8 percentile ranks according to each of the 8 metrics, and the scatterplot is constructed by plotting all pairs of percentile ranks. A diffuse scatterplot indicates low correlation, whilst a scatterplot concentrated along the diagonal indicates high correlation.

#### (1.1) Qualitative assessment and interpretation of metrics

We tabulate the best and worst performing navigation paths as evaluated by each of the 8 performance metrics (Figure 2 shows Level Appendix has Levels 6 and 11). Each row corresponds to a performance metric, the top 3 rows are the best-performing navigation paths, and the bottom 3 columns are the worst-performing navigation paths, according to the performance metric associated with each column. For example, the top 3 entries of the first column (labelled Dur) are the navigation paths that took the shortest time to complete, and the bottom 3 entries of the first column are the navigation paths that took the longest time to complete. Each navigation path is coloured according to the speed of navigation, with dark purple indicating slow navigation and bright yellow indicating fast navigation. The 3 checkpoints to visit are marked with white-blue circles.

We observe that the best navigators show similarly efficient navigation according to all 8 performance metrics, while the worst navigators show a curious diversity of behaviours. The most efficient navigation would be to directly visit each of the checkpoint in the shortest path connecting them, and these are precisely the navigation paths shown to be best according to all 8 performance metrics. Therefore the best-performing navigations *simultaneously* take a short time and length, have low curvature, stay far away from the map boundary, and follow popular navigation paths taken by many others. Meanwhile we observe a wide diversity of inefficient navigation in the worst-performers, indicating that there are qualitative differences in traits penalised by the 8 metrics. Firstly, we turn our attention to the extremely tangled navigation paths marked as the worst performers by the metrics Duration, Length, and M-Conformity. All of them exhibit a great deal of confusion in the player’s mind in figuring out what exactly to do with the game’s objective. The other worst-performers are not as markedly tangled, but we notice some traits more significantly picked up by other performance metrics, for example:

Row 5, Column 3: The player goes around many small loops in the beginning.
Row 5, Column 4: The player repeatedly bumps into the wall and has trouble steering back.
Row 6, Column 8: The player makes many abrupt pauses, marked by dark spots on the path.

#### (1.2) Quantitative comparisons between metrics

We study how the performance metrics are correlated. In Figure 3, the top row shows correlation coefficients for Levels 6, 8, and 11. Correlation coefficients are visualised with dark purple indicating low correlation and bright yellow indicating high correlation. In the bottom row, rank plots are shown; each square in the 8 x 8 tabulation shows a scatterplot of percentile ranks. For a pair of performance metrics X and Y, a rank scatterplot is produced as follows. For each one of the N=60,796 players in Level 6, let X_1_, … X_N_ be the rankings according to X and Y_1_, … Y_N_ be the rankings according to Y. Then we produce a scatterplot of N=60796 points by plotting the points (X_1_, Y_1_), … (X_N_, Y_N_). If X and Y are qualitatively similar, it would give similar rankings to the same path, making the points (X_1_, Y_1_), … (X_N_, Y_N_) concentrate near the diagonal. For example, when X=Y, all of the points will lie exactly on the diagonal, which is what we see on the 8 scatterplots that lie on the diagonal.

We observe that the 8 metrics show both consistency and diversity in assessing navigation paths. All correlation coefficients were found to be positive, ranging between 0.138 to 0.947 and with the average value of 0.605 ^2^. A more detailed look is made possible through rank plots. As seen in comparing one metric with itself, a perfect agreement between two metrics is indicated by all the points in a scatterplot lying along the diagonal. High correlation is similarly indicated by concentration of the scatterplot along the diagonal, and conversely low correlation is indicated by diffuse distribution of the scatterplot. Due to the 8 metrics being pairwise positively correlated whilst showing much diversity, we see that they are expected to enlarge the scope of behaviours captured.

### (2) Metrics are sensitive to navigation ability in the benchmark game players

We evaluate the effectiveness of metrics on the large benchmark dataset by splitting the dataset into 2 groups according to VOC, which stands for *visiting order correctness*. The VOC is a binary label, *correct* or *incorrect*, assigned to each navigation path indicating whether the numbered checkpoints were visited in the correct order, as they were instructed in the SHQ game. (See blue numbered circles in Figure 1) Since visiting checkpoints in the correct order is a simple task, we assert that VOC is an intuitively clear label for optimal path navigation. Among both males and females, 80.5% of the population had the *correct* visiting order for Level 6, 38.7% for Level 8 and 78.0% for Level 11. Here, the frequent failure in visiting checkpoints correctly in Level 8 can be attributed to many players missing the first turn to the left.

We found that players that visited checkpoints in the wrong order performed worse according to all 8 metrics. We further split the player groups into their ages (50 to 80), and examined distributions of scores given by the 8 performance metrics. As shown in Figure 4A, most metrics show a significant split across VOC groups, thus indicating that players of the incorrect visiting order (orange plot) perform worse than the players of the correct visiting order (blue plot). Each plot in Figure 4A shows the 25th, 50th, 75th percentile scores plotted across age, for Level 6. Each vertical slice is equivalent to a boxplot that marks 25th, 50th, and 75th percentile scores at a specific age. The solid line is for the 50th percentile and lightly shaded regions indicate the range between 25th and 75th percentiles. We note that the interquartile spreads between the two groups are particularly prominent for the metrics Duration, Length, M-Conformity and V-Conformity.

**Figure 4.**
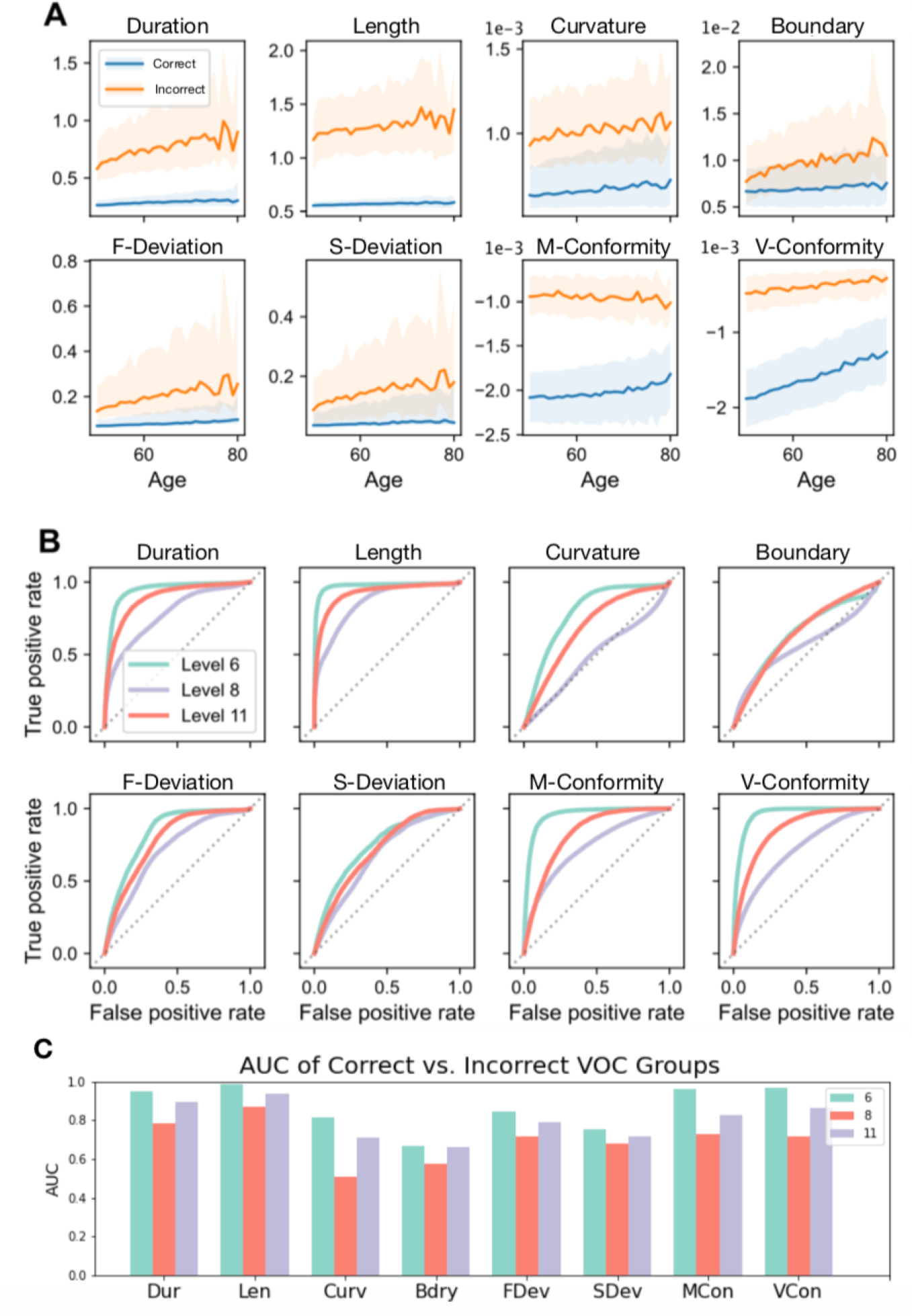
Sensitivity of the metrics to visiting order correctness (VOC). (A): Distributions of scores of the 8 metrics in Level 6, plotted across age (x-axis), for the correct VOC group (blue) and the incorrect VOC group (orange). Each vertical slice is equivalent to a boxplot that marks 25th, 50th, and 75th percentile scores at a specific age. The solid line is for the 50th percentile and lightly shaded regions indicate the range between 25th and 75th percentiles. (B): Receiver operating characteristic (ROC) curves for determining visiting order correctness, calculated for each of the 8 metrics and ROC curves for all 3 levels. Age is not incorporated here, unlike (A). (C): Area-under-curve (AUC) for the ROC curves in (B).

In Figure 4B, a series of receiver-operator characteristic (ROC) curves were computed to evaluate the sensitivity of the metrics to VOC. We compute ROC curves by using each metric as a scoring function to distinguish the Correct vs. Incorrect VOC groups. Area under the curve (AUC) values for these ROC curves are summarised in Figure 4C, and they indicate that the SHQ metrics are highly sensitive to performance in SHQ, with some variation between the three levels. The range of area-under-the-curve (AUC) computed for these ROC curves were 0.665-0.988 for Level 6, 0.507-0.872 for Level 8, and 0.664-0.934 for Level 11. In all three levels, the Length metric emerges as having the highest AUC value, whilst the Duration metric has the second-highest AUC value for Levels 8 and 11. The M-Conformity metric has the second highest AUC value for Level 6. Curvature and Boundary Affinity metrics generally show low AUC, although the Curvature metric still attains the AUC value >0.8 in Level 6. All of the AUC values were greater than 0.5. Similarly to AUC, t-tests were performed to distinguish the Correct vs. Incorrect VOC groups. The p-values obtained from the t-tests were all found to be <0.0005.

### (3) Metrics are sensitive to Alzheimer’s disease and APOE genotypes

We examine how the AD diagnosis and APOE genotype affects the 8 performance metrics. The *labelled dataset* used consists of the following cohorts: the AD patients (N=18), *APOE*ε3ε3 carriers (N=28), and *APOE*ε3ε4 carriers (N=31). Due to the small size of the labelled dataset, we assess performance of each navigation path by comparing the scores from the 8 performance metrics to those of the matched demographic from the large benchmark dataset (N=60,976), and computing the percentile of the score within the matched demographic^3^. Each percentile rank thus indicates relative performance of a navigation path, compared to others in the same demographic. The output of the comparison is displayed on Figure 5, where *medians* of percentile ranks in each cohort are used to create radar plots. Furthermore, t-tests were performed to distinguish the pair AD vs. *APOE*ε3ε3 and also the pair *APOE*ε3ε4 vs. *APOE*ε3ε3; the p-values are also recorded in Table 1.

**Table 1.**
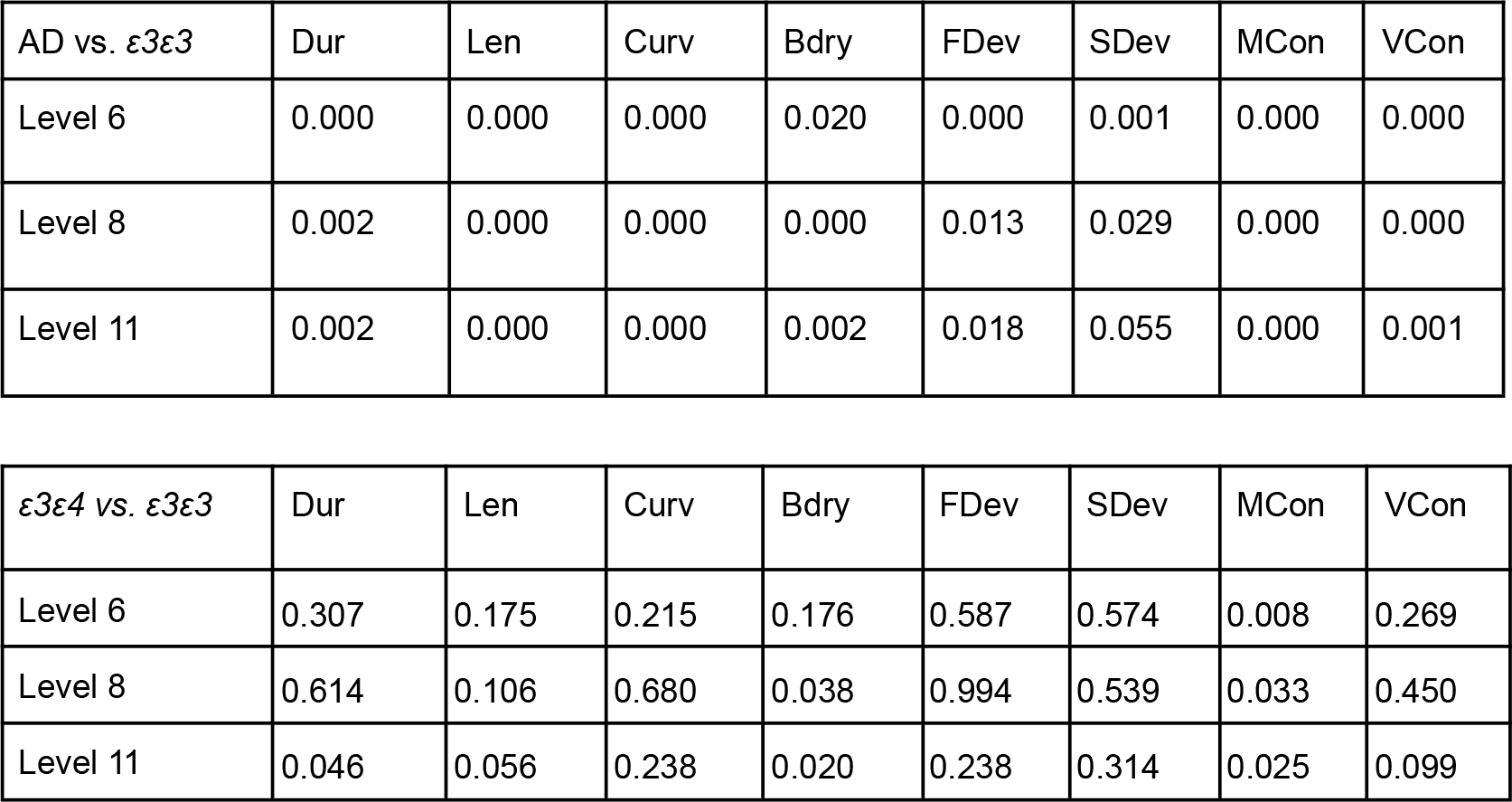
The t-test p-values obtained for distinguishing the AD cohort from the APOEε3ε3 cohort (top), and the APOEε3ε4 cohort from the APOEε3ε3 cohort (bottom).

**Figure 5.**
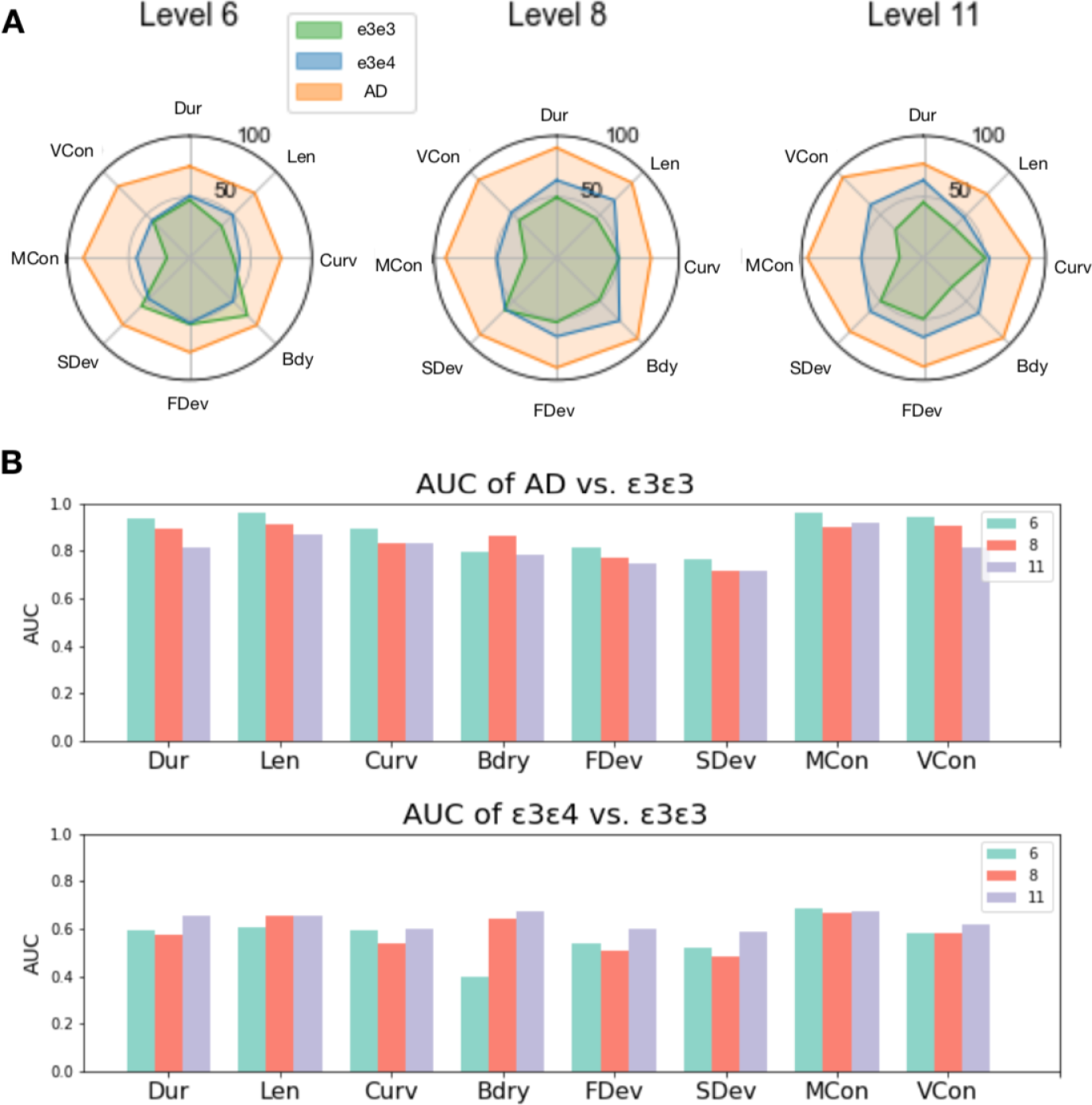
(A): Radar plots for the labelled dataset, across 3 levels (Levels 6, 8, 11), 8 performance metrics, and 3 labels (APOEε3ε3, APOEε3ε4, AD). Radar plots were constructed using medians of percentile ranks, where percentile rank was calculated by comparing performance of each navigation path to a matched group in the benchmark data. Note that higher percentile corresponds to worse performance, and that the poor performance of the AD group is easily noticeable from the large orange octagons. (B): AUC values obtained for distinguishing AD vs. APOEε3ε3 (top) and APOEε3ε4 vs. APOEε3ε3 (bottom).

We find that all of the 8 metrics show strong sensitivity to AD diagnosis, and show weak sensitivity to APOE genotypes (Figure 5). Here, high percentile corresponds to worse performance, and we immediately notice from the large orange octagons that the AD cohort performed poorly according to all 8 performance metrics, in all 3 gameplay levels. Crucially, we see that Curvature and Boundary affinity metrics that performed worse in distinguishing visiting order correctness (Figure 4) are highly sensitive to AD diagnosis, since they assign high percentiles to the AD cohort. In comparison to the uniformly poor performance detected from all 8 metrics, there is a weaker sensitivity detected from comparing the *APOE*ε3ε3 cohort to the genetically at-risk *APOE*ε3ε4 cohort. This is indicated from the green octagon (*APOE*ε3ε3) being slightly smaller than the blue octagon (*APOE*ε3ε4). This situation may be leveraged from carrying out the same analysis on a larger dataset, which will allow a more precise comparison between the two cohorts.

### (4) Metrics serve as early predictor of effective navigation

We study discriminatory power of the SHQ metrics on incomplete navigation paths. Here an “incomplete navigation path” is a fraction of the full path, which models situations when the player didn’t complete the gameplay challenge, quitting prematurely at some point of their navigation. This scenario is relevant for a diagnostic scenario where a patient playing the SHQ game for an AD diagnosis quits the game halfway, and performance assessment must be made with limited information. The wealth of information provided by our large benchmark data can be leveraged in this situation. We evaluate our metrics on navigation paths in the benchmark data that are *snipped* up to various lengths. This means that when tasked to assess the performance of an incomplete navigation path of length L, we can evaluate our metrics on the navigation path and compare these to scores in the benchmark data computed for navigation paths snipped up to length L.

We replace the Length metric by Velocity, which is just the average velocity of the navigation. This replacement is due to the fact that the Length metric loses meaning in our case when gameplay paths have been snipped up to a certain fixed length. The snipping length L has been chosen at the 0%, 20%, … 100% quantiles of the distribution of navigation lengths. Here the 100% quantile means the maximum of navigation lengths, so that snipping at 100% quantile produces no snipping at all.

Conversely, the 0% quantile means the minimum of navigation lengths, so that navigation lengths are snipped to the length L equal to the minimum of all navigation lengths. The choice to use quantiles is due to the distribution of navigation length being highly skewed: most navigation paths are short, and a tiny proportion of navigation paths are very long. The choice to use quantiles therefore directly references relative performance of the general populace. The values of AUC for telling apart visiting order correctness are tabulated in Figure 6, with one AUC value calculated for every pair of (Snip quantile, Performance metric). Here we use two colours to group the metrics, orange for Velocity and Duration metrics, and blue for the rest of the metrics, which are the new metrics proposed in this paper.^4^

**Figure 6.**
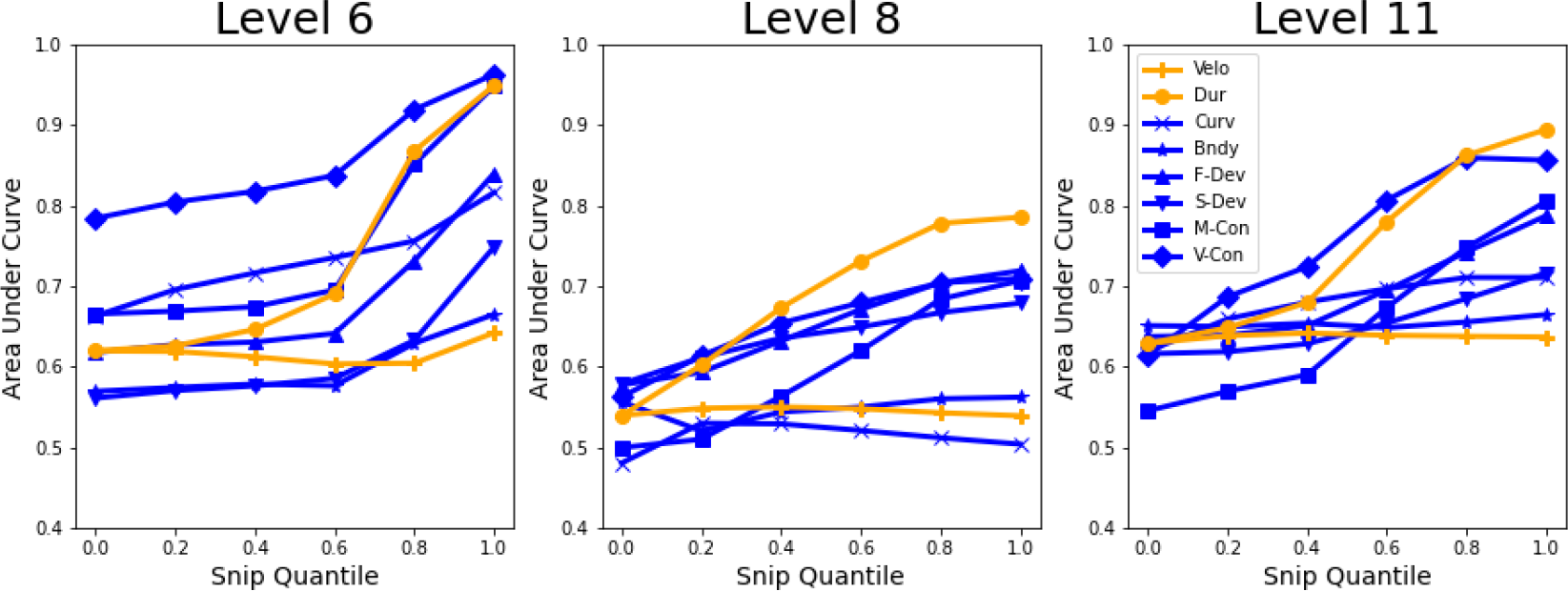
AUC values for discriminating visiting order correctness, calculated per each level, performance metric, and snip quantile. The snip quantile refers to the quantile in the distribution of navigation lengths in each level. Snip quantile = 1.0 means navigation paths are left untouched, whilst snip quantile = 0.0 means navigation paths are snipped to the minimum length among all paths. Metrics in yellow are those used in past analysis. Metrics in blue are the 6 proposed performance metrics.^5^

We find a situation where several of the proposed metrics are superior to the existing ones when working with incomplete navigation paths. This is indicated in small snip quantiles 0%, 20%, 40% of Level 6, in which Curvature, M-Conformity and V-Conformity stay above the Duration metric which otherwise shows great performance. In general, V-Conformity shows comparably strong performance to the Duration metric, yet in Level 6 the performance is markedly better for small snip quantiles. Thus in Level 6, conformity to the population behaviour, measured using V-Conformity, is a better *early predictor* than fastness for whether a player will visit the checkpoints in the correct order. Further analysis with other maps will illuminate the property of a map that contributes to V-Conformity being particularly effective here. We additionally remark that the general rising trend in all plots is in line with the principle that more information of navigation path should provide more accurate diagnostic information, and at snip quantile 100%, gameplay paths are untouched and we recover the previous analysis in Figure 4.

## DISCUSSION

Our results show that mathematically refined analysis of spatial navigation offers a larger toolbox to understand path navigation. We introduce 6 novel performance metrics and apply them to the SHQ game, capturing diverse properties of human navigation behaviour. The proposed new performance metrics assess curviness, affinity to boundary, and deviation / conformity to the population behaviour. The conformity and deviation from benchmark behaviour were measured in four diverse ways arising from the concept of the origin-destination matrix. By examining best-performers, worst-performers and inter-metric correlations, we find that the performance metrics capture a wide range of qualitative behaviours whilst all being positively correlated.

Performance metrics were validated by partitioning navigation paths into two groups, depending on whether the checkpoints were visited in the correct order. Here, the Length metric emerged as the strongest signal for distinguishing the players with suboptimal navigation. The Duration, M-Conformity, and V-Conformity metrics also showed significant AUC value of >0.7. The Curvature and Boundary Affinity metrics did not show as significant AUC values, although the AUC value for the Curvature metric in Level 6 was >0.8. All AUC values were greater than 0.5.

Consistent with previous evidence of spatial disorientation in AD (Levine et al., 2020; Lithfous et al., 2013; Tuena et al., 2021; Vlček, K., & Laczó, 2014), the metrics showed high sensitivity for AD diagnosis, all 8 performance metrics having AUC >0.7 in all 3 levels according to the rank percentile distribution when compared to the *APOE*ε3ε3 carriers. Here, the Length metric again emerges as the strongest indicator for the AD patients, although the M-Conformity and V-Conformity metrics showed comparably high AUC values. Taking into account the length/duration-normalisations performed in the 6 new performance metrics, this analysis is in agreement with the expected behaviour of curvy, boundary-seeking (Hardcastle et al., 2015), navigation in AD patients.

Genetic variation in the *APOE* gene is currently the strongest known genetic risk factor for sporadic AD Compared with the ε3ε3 carriers, those with the ε3ε4 show a threefold to fourfold increased risk for AD (Farrer, 1997; Liu et al., 2013). The mid-life *APOE* ε4 carriers AD showed subtle differences relative to the benchmark cohort, with M-Conformity emerging as the metric exhibiting the strongest signal of distinguishing the ε3ε4 carriers from the ε3ε3 carriers. Boundary affinity exhibited sensitivity to *APOE* carriers in Levels 8 and 11, consistent with previous research (Bierbrauer et al., 2019; Hardcastle et al., 2015, Coughlan et al., 2019, Coughlan et al., 2020).

We find that our proposed metrics show much promise in assessing path navigation online, when we calculate the metrics at each moment during a gameplay. The proposed gameplay metrics either show comparable or superior performance to the baseline metrics (Length and Duration), as shown on Figure 6, especially for Level 6 where the V-Conformity metric significantly outperforms the Duration metric when gameplay paths have been snipped to short lengths. The proposed online evaluation opens the possibility to deploy the metrics when patients undergoing AD assessment quit halfway due to frustration or fatigue, but also in other types of mobility datasets where the endpoint of a trajectory is not necessarily known.

Our finding was limited by the small sample size for the cohorts labelled with AD diagnosis and *APOE* alleles, making it necessary for the findings to be replicated in larger samples in the future. Comparing distributions of metrics among the benchmark cohorts of correct and incorrect visiting order (Figure 4) could be repeated for players with and without AD, resulting in a direct comparison of navigation abilities of those with and without AD. In addition, there was a limited analysis of the interaction between the geometry of SHQ levels and player navigation. Apart from the increasingly complicated terrain as one progresses from Levels 6, 8, and then 11, it is hard to make a definitive conclusion on how players respond to a wider diversity of terrain geometry. Future studies may complement this by studying navigation on levels that become increasingly challenging in specific ways, such as having more checkpoints or having more curvy terrains.

In conclusion, the proposed performance metrics have a potential to be a comprehensive pre-clinical screening toolbox for Alzheimer’s Disease. Prevalence of mobile computing devices makes remote diagnosis through navigation especially viable. A more careful analysis involving a larger dataset of clinically labelled navigations is necessary to achieve this. The comprehensive behavioural traits captured by our metrics, combined with their efficiency in an on-line context, make them particularly promising to study ethological data sets, for instance based on GPS, where they could be used to quantify navigation of birds, insects, and people (Ghosh et al. 2022, Torus et al. 2017, Thums et al. 2018).

## MATERIALS AND METHODS

### Participants

Two populations of SHQ game players were analysed: benchmark and clinical label. The benchmark population was collected via the Apple and Android app store, with demographic information but without any clinical labels. The clinical population included demographic information and *APOE* genotype status / AD clinical diagnosis.

### Benchmark population (N = 60,976)

A unique population-level benchmark navigation data was generated by extracting a subset of the global SHQ database that matched the demographic profile of the clinical label cohorts, namely male and female players from the United Kingdom aged 50–80 years old (Table 1). This consisted of 60,976 adults (female-to-male ratio 52/48) which we considered to be a representation of navigation behaviour in the general population. We analysed in total just under 161,000 unique navigation paths across three levels in SHQ. Participants were given the option to opt in or opt out of the data collection when they played the game on their personal mobile phone, iPad, or tablet. If a participants’ response were to opt in, their SHQ data were anonymized and stored securely by the T-Systems’ data centre under the regulation of German data security law. Ethical approval was previously granted by the Ethics Research Committee CPB/2013/015.

### At-risk and clinical AD cohort (N = 77)

The at-risk and clinical AD cohorts consist of 77 individuals further subdivided into 3 subgroups, corresponding to players with: clinical Alzheimer’s disease diagnosis (N=18), cognitively normal *APOE*-ε3ε3 (N=28) and *APOE*-ε3ε4 individuals (N=31). The ε3ε4 individuals are at heightened genetic risk of AD compared to the ε3ε3 individuals (Table 1).

The clinical groups were prescreened for a history of psychiatric or neurological disease, history of substance dependence disorder, or any significant relevant comorbidity. All participants had normal or corrected-to-normal vision. Early-stage Alzheimer’s disease patients from the community were recruited via The Dementia Research and Care Clinic study. Clinical diagnosis was made by a consultant at the Norfolk and Suffolk Foundation Trust by interviewing the patient, examining neuropsychological assessment scores, structural clinical MRI scans, and the patient’s medical history which met the diagnostic criteria (McKhann et al., 2011). For the genetically-at-risk *APOE* genotype participants, saliva samples were collected from those who passed this screening. See (Coughlan et al., 2019) for more information on the genotyping procedure. Only ε3ε3/ε3ε4 variants of the *APOE* genotype were retained as *APOE*ε3ε4 is known to be one of the biggest risk factors for developing symptomatic AD. ε3ε4 carriers (23% of the global population) show a threefold to fourfold increased risk, compared to ε3ε3 carriers (75% of the population). Written consent was obtained from all participants, and ethical approval was obtained from Faculty of Medicine and Health Sciences Ethics Committee at the University of East Anglia (reference FMH/2016/2017-11) and from the United Kingdom National Research Ethics Service (16/LO/1366).

### The SHQ Game

In SHQ, the player embodies the son of an old sea adventurer, whose memories of mythical creatures faded over time due to Alzheimer’s Disease. To help recover his memories, the player adventures on a boat to find his father’s old notes. We focus on the gameplay challenge known as wayfinding. In wayfinding levels, the player is first provided with a map with the outline of the shore, the starting position of the boat, and the location of a few checkpoints with numerical labels: 1,2, and so on (see Figure 1). After a while, the map is hidden from the player and the player must navigate a virtual terrain described by the map to visit checkpoints in the correct order. In this research, we focus on analysing the navigation paths in these wayfinding levels, assessing how well each player completes each level using various performance metrics. A navigation path is stored as a list of coordinates in a discrete two-dimensional grid. The positions are sampled at a constant rate of 2 Hz. Due to this low sampling rate and the coarse grid, the navigation paths originally recorded are crude in resolution (Figure 1, Panel B). Nevertheless, convincing estimates of true game-play paths underlying the crude sampling can be recovered using Gaussian kernel smoothing (Figure 1, Panel C).

### Performance Metrics

We introduce 6 new performance metrics that expand the 2 previously used metrics of Duration and Length (Coughlan et al., 2019; Coutrot et al., 2018). In total, the 8 performance metrics are: (1) Duration, (2) Length, (3) Curvature, (4) Boundary affinity, (5) F-Deviation, (6) S-Deviation, (7) M-Conformity, and (8) V-Conformity. The qualitative meaning behind them are as follows:

(1) Duration: How long a player took to complete the level
(2) Length: The length of the completed navigation path
(3) Curvature: Length-averaged curvature
(4) Boundary affinity: Length-averaged closeness to the boundary
(5) F-Deviation and (6) S-deviation: Time-averaged deviation from benchmark navigation behaviour
(7) M-Conformity and (8) V-Conformity: Time-averaged conformity to the benchmark navigation behaviour

Below are details of the precise mathematical definitions of the proposed metrics. The *curvature* metric of a navigation path *γ* : [0, *T*] → ℝ^2^ is given by:

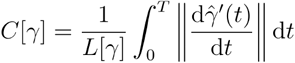

where *γ′*(*t*) = d*γ*/d*t*., 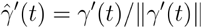, and we performed length-normalisation:

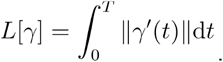

The *boundary affinity* metric of a navigation path *γ* : [0, *T*] → ℝ^2^ is given as the length-average of the closeness function *β*:

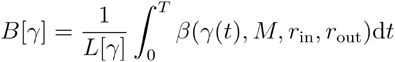

The closeness function (*βx, M, r*_*in*_, *r*_*our*_) uses the sigmoid function f(*t*) = 1/(1 + e^−t^) to compute the closeness of a point to the set which represents land mass in SHQ that the player cannot steer to. It assigns the value nearly 1 at distance *r*_in_ and the value nearly 0 at distance *r*_out_ from *M*:

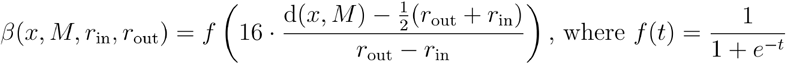

The metrics (5) (6) (7) (8) all represent the deviation of each navigation path against the benchmark population. As such, we may refer to these as *relative* whereas we may refer to (1) (2) (3) (4) as *absolute*, since they do not depend on other navigation paths. These *relative* metrics are all defined using the *origin-destination (OD) matrix*, a comprehensive summary of the population-level information of instantaneous movements. We also remark that the metric (8) is defined using cruder information than the OD matrix, the *V-Conformity*. The V-Conformity summarises the average direction emanating from each map coordinate.

The following is a mathematically precise description of the relative metrics. The OD matrix OD(Γ) is defined for each collection of paths Γ = {*γ*_1_ … *γ*_*N*_}. Let S ={*p*_1_ … *p*_*m*_} ⊆ ℝ^2^ be a set of points that each *γ*_*i*_ takes value in, so that each *γ*_*i*_ is a function *γ*_*i*_ : {0,1,…*T*_*i*_ -1}→*S*. Now given two points *P*_*i*_ and *P*_*j*_, we can count the frequency at which paths in Γ transition from *P*_*i*_ to *P*_*j*_ by counting all pairs (*γk*,*t*) of paths *γk* and a time *t* such that *γk*(*t*) = *Pi* and *γk* (*t* +1) = *P*_*i*_. This is precisely how we define the OD matrix: OD(Γ) is a *m* × *m* matrix whose (*i,j*) th entry is defined as:

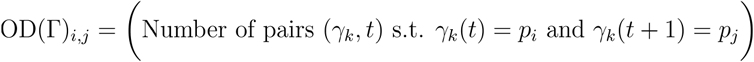

Now let *γ*:{0,1⃛ *T* -1} →*S* be a navigation path we would like to assess, let Γ = {*γ*_1_,⃛ *γ*_*N*_} be the navigation paths of the benchmark population, and let *S* be the collection of points on the map, sampled on a discrete square grid. Then the (5) F-Deviation metric and (6) S-Deviation metric are defined by comparing the OD matrix of the singleton set {*γ*} versus the OD matrix of Γ :

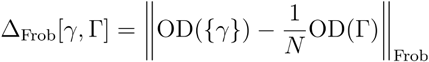

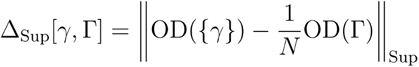

Here, the meaning of the terms Frobenius and supremum become clear - they refer to the way in which the matrix norm is taken (the supremum norm is also known as the *L*^∞^ norm).

The *M-Deviation* metric is defined by summing all the entries on OD(Γ) that correspond to nonzero entries of OD({*γ*}),

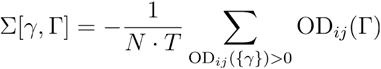

Intuitively, the sum will be large when the path *γ* follows a “popular” path that is commonly found in the normative data Γ which is used for comparison. The minus sign reverses the order so that goes through common locations is scored lower than a path that does not.

While the matching-sum metric captures what are typical paths, we are also interested in the average at each location. Similarly to (Mazzoli et al. 2019), we define the *mobility field* F over the coordinate set *S* by summing all the transitions happening at a location across a single time step and normalising it.

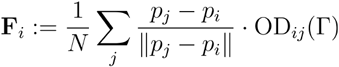

The last relative metric which we call the *V-Conformity* of a navigation path *γ* is then defined by taking the “path integral” of the path through the mobility field:

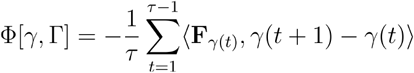

where **F**_*γ*(*t*)_ is the mobility field at *γ*(*t*), defined by **F**_*γ*(*t*)_ = **F**_*i*_ where . As before, the minus sign makes the mobility functional large when a navigation path follows closely the most common transitions recorded on the V-Conformity **F**.

To furnish a sensible comparison, the set of navigation paths to compare against were chosen to be near each player’s age range. To be precise, given a player of age n, the four relative performance metrics were computed by comparison against the benchmark population of age *n* - 5 to *n* + 5, with appropriate weighting.

### Normalisation

All performance metric scores appearing in the analysis above were normalised by comparison with Levels 1 and 2 of SHQ. More precisely, for each player that scored X in some performance metric, the normalised score is defined by X/(L_1_ + L_2_), where L_1_ and L_2_ are the lengths of the navigation of the player in Levels 1 and 2.

### Statistical analysis

The data were analysed using Python3, version 3.8.1. For each level, a percentile score was computed. This percentile score reflected the relative performance between the age and sex matched benchmark, and each clinically labelled participant. Higher percentile values reflect poorer performance. Only benchmark players who visited the checkpoints correctly were used in the normative sample to compare the clinical label population against. Receiver operating characteristic (ROC) curves quantified the sensitivity of the metrics to predict visiting order correctness in the benchmark navigation data and clinical labels in the clinical population.

## Data and code availability

A dataset with the preprocessed trajectory lengths and demographic information is available at https://osf.io/7nqw6/?view_only=6af022f2a7064d4d8a7e586913a1f157. Owing to its considerable size (around 1 terabyte), the dataset with the full trajectories is available on a dedicated server: https://shqdata.z6.web.core.windows.net/. Future publications based on this dataset should add ‘Sea Hero Quest Project’ as a co-author.

The Python and MATLAB (R2018a) code that allows the presented analyses to be reproduced is available along the preprocessed trajectory lengths and demographic information at https://osf.io/7nqw6/?view_only=6af022f2a7064d4d8a7e586913a1f157.

## Data Availability

A dataset with the preprocessed trajectory lengths and demographic information is available at https://osf.io/7nqw6/?view_only=6af022f2a7064d4d8a7e586913a1f157. Owing to its considerable size (around 1 terabyte), the dataset with the full trajectories is available on a dedicated server: https://shqdata.z6.web.core.windows.net/. Future publications based on this dataset should add 'Sea Hero Quest Project' as a co-author.
The Python and MATLAB (R2018a) code that allows the presented analyses to be reproduced is available along the preprocessed trajectory lengths and demographic information at https://osf.io/7nqw6/?view_only=6af022f2a7064d4d8a7e586913a1f157.

## ACKNOWLEDGEMENTS

U.L. and H.A.H. are members of the UK Centre for Topological Data Analysis supported by EPSRC grant EP/R018472/1.

U.L. is supported by the Korea Foundation for Advanced Studies.

R.L-C and H.A.H thank St John’s College Oxford for research funding support.

H.A.H. gratefully acknowledges funding from EPSRC EP/R005125/1 and EP/T001968/1, the Royal Society RGF\EA\201074 and UF150238.

R.L. acknowledges support from the EPSRC Grants EP/V013068/1 and EP/V03474X/1.

G.C. is supported by Alzheimer’s Association (AARF-23-1151259) and Alzheimer Society of Canada (22-08)

M.H. is supported by MRC IAA, MRC CARP, RST INDICATE, NHMRC SNAVI.

H.S. is supported by Alzheimer’s Research UK (ARUK-DT2016-1).

## CONFLICT OF INTEREST

We declare no conflict of interest.

## APPENDIX

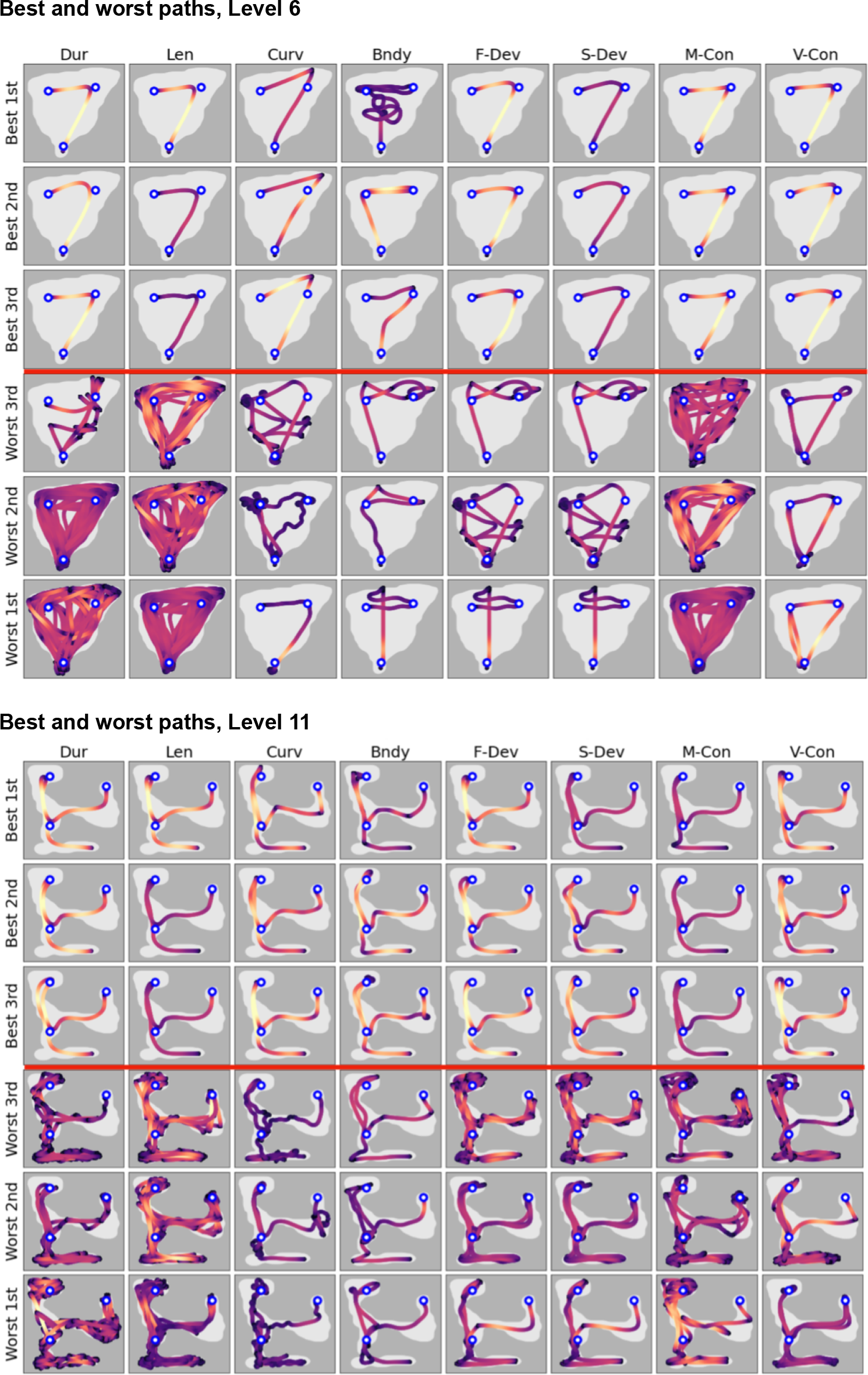

**Table 2:** Sample size and gender ratio of populations studied.

| | Benchmark | Alzheimer's disease | APOEε3ε3 | APOEε3ε4 |
| --- | --- | --- | --- | --- |
| Number of Players |  |  |  |  |
| Level 6 | 60,976 | 18 | 28 | 31 |
| Level 8 | 54,832 | 15 | 28 | 31 |
| Level 11 | 45,113 | 11 | 28 | 31 |
| Gender ratio (F/M) |  |  |  |  |
| Level 6 | 52/48 | 56/44 | 54/46 | 35/65 |
| Level 8 | 51/49 | 47/53 | 54/46 | 35/65 |
| Level 11 | 50/50 | 45/55 | 54/46 | 35/65 |

**Table 3.** AUC and t-test p-values, for distinguishing Correct vs. Incorrect VOC groups (Benchmark data)

| **SHQ Level 6** |  |  |  |  |  |
| --- | --- | --- | --- | --- | --- |
| *Metrics* | *AUC* | *Standard error* | *95% CI* |  | *t-test* |
|  |  |  | *Lower* | *Upper* | *P value* |
| Duration | 0.949 | 0.001 | 0.947 | 0.951 | 0.000 |
| Length | 0.988 | 0.000 | 0.987 | 0.988 | 0.000 |
| Curvature | 0.816 | 0.002 | 0.813 | 0.820 | 0.000 |
| Boundary affinity | 0.665 | 0.003 | 0.659 | 0.670 | 0.000 |
| Frobenius deviation | 0.842 | 0.002 | 0.839 | 0.846 | 0.000 |
| Supremum deviation | 0.750 | 0.002 | 0.745 | 0.755 | 0.000 |
| M-Conformity | 0.964 | 0.001 | 0.963 | 0.966 | 0.000 |
| Mobility functional | 0.965 | 0.001 | 0.963 | 0.966 | 0.000 |
| **SHQ Level 8** |  |  |  |  |  |
| *Metrics* | *AUC* | *Standard error* | *95% CI* |  | *t-test* |
|  |  |  |  |  | *P value* |
| Duration | 0.786 | 0.002 | 0.782 | 0.790 | 0.000 |
| Length | 0.872 | 0.002 | 0.869 | 0.875 | 0.000 |
| Curvature | 0.507 | 0.003 | 0.502 | 0.511 | 0.000 |
| Boundary affinity | 0.573 | 0.002 | 0.568 | 0.578 | 0.000 |
| Frobenius deviation | 0.718 | 0.002 | 0.714 | 0.723 | 0.000 |
| Supremum deviation | 0.680 | 0.002 | 0.675 | 0.684 | 0.000 |
| M-Conformity | 0.731 | 0.002 | 0.727 | 0.735 | 0.000 |
| Mobility functional | 0.718 | 0.002 | 0.714 | 0.722 | 0.000 |
| **SHQ Level 11** |  |  |  |  |  |
| *Metrics* | *AUC* | *Standard error* | *95% CI* |  | *t-test* |
|  |  |  |  |  | *P value* |
| Duration | 0.894 | 0.002 | 0.891 | 0.898 | 0.000 |
| Length | 0.934 | 0.001 | 0.932 | 0.937 | 0.000 |
| Curvature | 0.710 | 0.003 | 0.705 | 0.715 | 0.000 |
| Boundary affinity | 0.664 | 0.003 | 0.659 | 0.670 | 0.000 |
| Frobenius deviation | 0.787 | 0.002 | 0.782 | 0.791 | 0.000 |
| Supremum deviation | 0.718 | 0.003 | 0.712 | 0.723 | 0.000 |
| M-Conformity | 0.829 | 0.002 | 0.825 | 0.833 | 0.000 |
| Mobility functional | 0.862 | 0.002 | 0.858 | 0.865 | 0.000 |

**Table 4.**
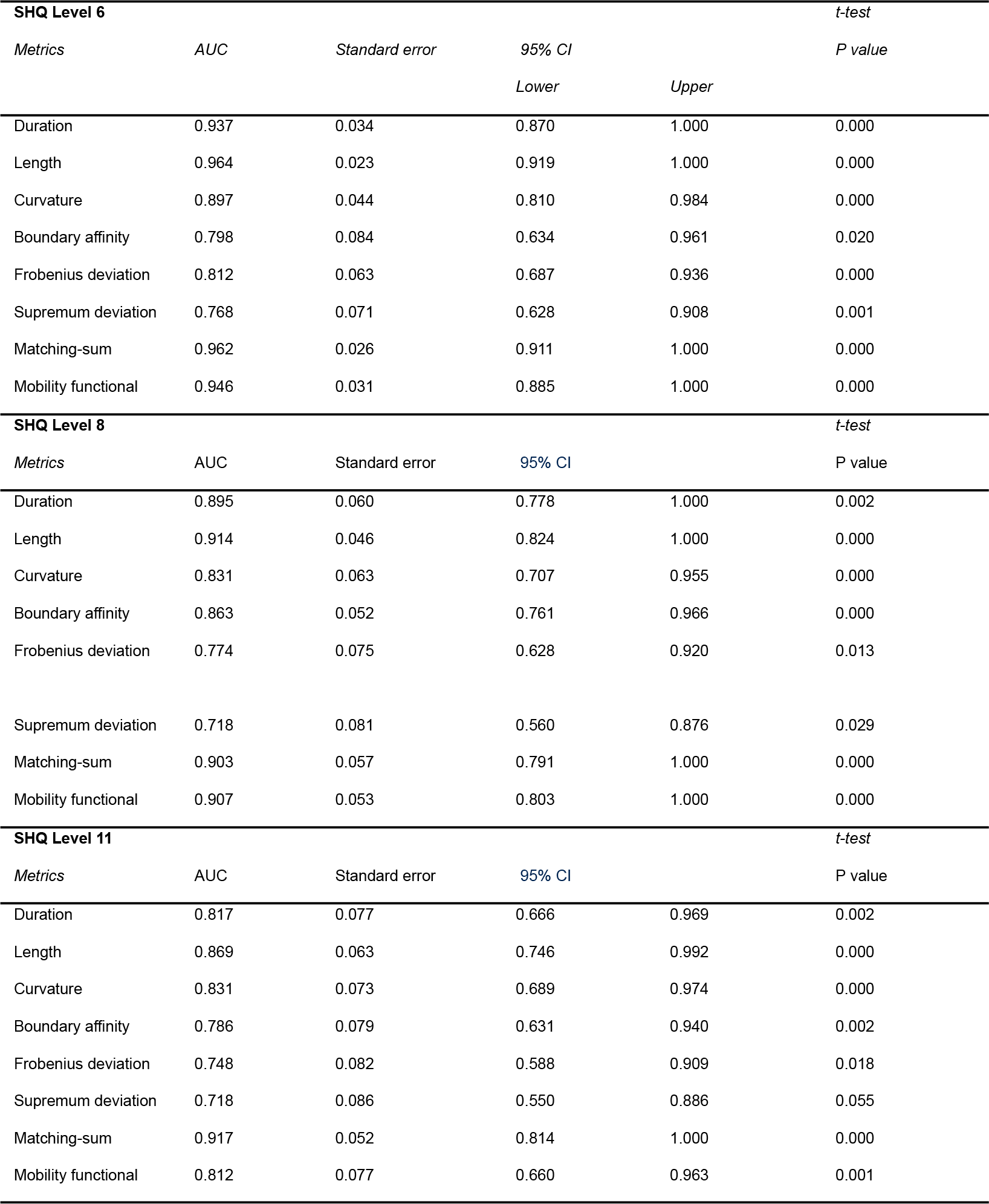
AUC and t-test p-values, for distinguishing AD vs. APOEε3ε3 groups.

**Table 5.** AUC and t-test p-values, for distinguishing APOEε3ε4 vs. APOEε3ε3 groups.

| SHQ Level 6 |  |  |  |  |  |
| --- | --- | --- | --- | --- | --- |
| Metrics | AUC | Standard error | 95% CI |  | t-test |
|  |  |  | Lower | Upper | P value |
| Duration | 0.591 | 0.076 | 0.443 | 0.739 | 0.307 |
| Length | 0.605 | 0.075 | 0.458 | 0.752 | 0.175 |
| Curvature | 0.596 | 0.076 | 0.447 | 0.744 | 0.215 |
| Boundary affinity | 0.395 | 0.076 | 0.246 | 0.544 | 0.176 |
| Frobenius deviation | 0.536 | 0.078 | 0.383 | 0.688 | 0.587 |
| Supremum deviation | 0.522 | 0.079 | 0.367 | 0.676 | 0.574 |
| Matching-sum | 0.685 | 0.071 | 0.547 | 0.824 | 0.008 |
| Mobility functional | 0.584 | 0.076 | 0.435 | 0.733 | 0.269 |
| SHQ Level 8 |  |  |  |  | t-test |
| Metrics | AUC | Standard error | 95% CI |  | P value |
| Duration | 0.578 | 0.076 | 0.430 | 0.727 | 0.614 |
| Length | 0.653 | 0.072 | 0.511 | 0.795 | 0.106 |
| Curvature | 0.540 | 0.077 | 0.389 | 0.692 | 0.680 |
| Boundary affinity | 0.645 | 0.074 | 0.500 | 0.790 | 0.038 |
| Frobenius deviation | 0.510 | 0.077 | 0.360 | 0.661 | 0.994 |
| Supremum deviation | 0.486 | 0.078 | 0.333 | 0.639 | 0.539 |
| Matching-sum | 0.669 | 0.071 | 0.531 | 0.803 | 0.033 |
| Mobility functional | 0.582 | 0.076 | 0.434 | 0.730 | 0.450 |
| SHQ Level 11 |  |  |  |  | t-test |
| Metrics | AUC | Standard error | 95% CI |  | P value |
| Duration | 0.657 | 0.074 | 0.512 | 0.801 | 0.046 |
| Length | 0.656 | 0.074 | 0.511 | 0.800 | 0.056 |
| Curvature | 0.600 | 0.075 | 0.454 | 0.747 | 0.238 |
| Boundary affinity | 0.675 | 0.071 | 0.536 | 0.813 | 0.020 |
| Frobenius deviation | 0.597 | 0.076 | 0.449 | 0.745 | 0.238 |
| Supremum deviation | 0.586 | 0.075 | 0.439 | 0.734 | 0.314 |
| Matching-sum | 0.675 | 0.071 | 0.536 | 0.815 | 0.025 |
| Mobility functional | 0.618 | 0.076 | 0.470 | 0.767 | 0.099 |

## Footnotes

1 Boundary affinity has been linked to temporal grid cell-like representation in the entorhinal cortex of *APOE* ε4 carriers are functionally unstable, leading to a boundary-driven error correction during wayfinding (Bierbrauer et al., 2019; Hardcastle et al., 2015, Coughlan et al., 2019, Coughlan et al., 2020).

2 The minimum, maximum, average correlation coefficients for each level are the following. For Level 6, it is (0.249, 0.911, 0.59). For Level 8, it is (0.138, 0.922, 0.595). For Level 11, it is (0.175, 0.947, 0.629).

3 Matched demographic is defined to be players in the Benchmark dataset of the same age and gender.

4 Performances were evaluated for a randomly chosen 50% of the benchmark data, where the other 50% was used to compute the origin-destination matrix representing population behaviour in the Deviation and Conformity metrics. This is an analogous process to the Test-Train split in machine learning literature, to make sure that the test data was not used for training a classifier.

5 The following is a detailed description of how navigation path snipping works for benchmark dataset. As before, navigation paths in the benchmark dataset are divided into 2 groups, depending on whether the player visited checkpoints correctly or not. Fix a length L and snip each navigation path up to length L, starting from the beginning. If a navigation path had length less than L, then that navigation path is left untouched. Each navigation path is then assessed with the proposed performance metrics, but computed for the incomplete navigation path of length ≤ L, instead of using the full complete navigation path. With this data, we can run the ROC curve analysis as before and obtain the AUC value. The AUC allows us to evaluate whether the performance metric was effective.

## Notes

### Competing Interest Statement

The authors have declared no competing interest.

### Author Declarations

Ethics Research Committee of the Department of Cognitive, Perceptual and Brain Sciences of University College London (CPB/2013/015) gave ethical approval for the benchmark population data in this work. Faculty of Medicine and Health Sciences Ethics Committee at the University of East Anglia (reference FMH/2016/2017-11) and United Kingdom National Research Ethics Service (16/LO/1366) gave ethical approval for the clinical and at-risk population data in this work. Written consent was obtained from all participants for this dataset.

